# Investigation and analysis of the clinical application of Perampanel in the Southeast of China

**DOI:** 10.1101/2023.04.04.23288128

**Authors:** Lingqi Li, Yan Huang, Lei Ma, Zhiyu Shao

**Affiliations:** The pharmacy department of the First Affiliated Hospital of Xiamen University, Xiamen, 361102, China; The Department of Neurology of the First Affiliated Hospital of Xiamen University, Xiamen, 361102, China; Department of Pharmacy, The First Affiliated Hospital of Xiamen University, Xiamen, 361005, China

## Abstract

**Objective:** The aim of this study was to investigate the clinical trials of Perampanel (PER) at the First Affiliated Hospital of Xiamen University (FAHXU). Recommendations have been developed for the use of Perampanel in clinical practice.

**Design:** A retrospective single-center validation study.

**Setting:** A tertiary general hospital in Xiamen, a city located on the southeastern coast of China.

**Participants:** A total of 831 prescriptions were included for 188 patients were treated with Perampanel.

**Outcome measures:** Record the patient’s medication, including the diagnosis of the patient’s disease, the dose, course of medication, and the combination of medication, analyze the characteristics of the drug population and the medication unreasonableness by comparing the drug instructions and referring to relevant literature and guidelines.

**Results:** PER was mainly used in Neurology. Epilepsy is the most common conditions prescribed. There were 119 cases of inconsistent drug indications, 56 inappropriate dosing frequency, 3 inconsistent dose, and 6 inconsistent dose adjustment intervals.

**Conclusion:** There is an improper use of PER within the FAHXU. Greater attention should be paid to changes in patients’ condition and observation of discomfort following medication administration. Monitoring of PER use should be further enhanced. A rational drug evaluation system should be established to promote the rational use of PER.

**Article Summary:** Our study was one of the few studies on the clinical use of Perampanel, especially in Southeast China.

As a third-generation antiepileptic drug, data on the clinical rational drug use of Perampanel is limited in China.

There were several limitations worth noting. Firstly, the data in our study were collected at only one institution, thus not covering a broader population from other regions. Clinical outcomes, including medication efficacy and adverse events, were not tracked in patients with irrational medication use.

**Strengths and limitations:** Our study was one of the few studies on the clinical use of Perampanel, especially in Southeast China. As a third-generation antiepileptic drug, data on the clinical rational drug use of Perampanel is limited in China.

There were several limitations worth noting. Firstly, the data in our study were collected at only two institutions, thus not covering a broader population from other regions. Clinical outcomes, including medication efficacy and adverse events, were not tracked in patients with irrational medication use.

## Introduction

Epilepsy is a chronic, noncommunicable disorder of the brain characterized by transient central nervous system dysfunction^1^. Seizure are defined as abnormal neuronal discharges in the brain^2^. It has paroxysmal, transitory, repetitive, and stereotypic features^3, 4^. The symptoms of epilepsy are usually body convulsions and tremors, with more severe episodes of momentary loss of consciousness or abnormal sensations lasting seconds to minutes^5^. The global incidence of epilepsy is approximately 50 per 10,000 people, with at least 50∼70 million people are afflicted, indicating Epilepsy is one of the most common brain disorders ^6, 7^. With 80% of epilepsy patients residing in developing countries^8^. The recurrence of seizures has a serious impact on the patient’s physical and mental health and impose a significant burden on the family and society^9^. The management of epilepsy is therefore an important and pressing clinical issue. In general, there are many methods to treat illness and relieve symptoms, including diet therapies, medication, surgery, psychotherapy and immunotherapy^10^. The main tool to control seizures is drugs^11^.

Antiepileptic drugs (AEDs) can be roughly divided into three generations based on listing time. Valproic Acid, Ethyl Succinate, and Phenytoin are some of the first-generation AEDs. Oxcarbazepine, Levetiracetam and Topiramate are the second-generation AEDs. Lacamide, Isodoxorubicin Acetate and Perampanel represent the third generation of antiepileptic medications^12, 13^. Perampanel (PER) (Fycompa^®^) is developed by the Japanese pharmaceutical company Eisai. As an new anti-epileptic drug, PER is a highly selective and non-competitive antagonist of alpha-amino-3-hydroxy-5-methyl-4-isoxazole-propionic acid (AMPA) glutamate receptors^14, 15^.

PER, by targeting inhibition of postsynaptic AMPA receptors, induces an increase intracellular calcium ion, thus reducing neuronal excitability^16^. The mechanism of PER is distinct from that of other antiepileptic drugs, opening a new avenue for the treatment of epilepsy. PER was launched as a new antiepileptic drug in China in July 2021. As of January 2022, it is used at FAHXU. There are nearly 10 million patients with epilepsy in China, with approximately 400,000 new cases each year^17^. Relatively little evidence exists to guide PER use in China. The purpose of this article is to present an analysis of the unreasonable situation of PER in FAHXU in order to provide a reference for clinically irrational drug use, with the goal of promoting rational drug use, so that this drug can exert its best effect in the case of rational drug taking.

## METHODS

### Study design, data source and patient population

We used the Hospital Information System (HIS) at FAHXU, to collect PER prescription data from January 2022 to January 2023. FAHXU is a famous tertiary hospital in Southeast China, with 4,500 staffs, 2500 beds, and 5 million ambulatory care visits annually. It consists of 65 departments and treats patients of all ages. There were 100,000 visits to the neurology department each year. These data include patient department, basic physiologic information, drug use and dosage, medication duration, and concomitant medications. And compare the drug instructions, refer to relevant literature and guidelines, and analyze the rationality of the characteristics of the drug users and the phenomenon of irrational drug use. In order to analyze the rationality of the PER use situation in FAHXU, we also compared the PER package insert.

## RESULTS

From January 2022 to January 2023, PER was prescribed to a total of 188 patients in FAHXU, with a total of 831 prescriptions. Table 1 shows the basic characteristics of patients treated with PER, including gender, age and Body Mass Index (BMI).

**Table 1.** Demographic and clinical characteristics of epilepsy patients

| Patients' basic situation | Copies | Proportion | Patients' basic situation | Copies | Proportion |
| --- | --- | --- | --- | --- | --- |
| Male | 111 | 59.04% | < 12 years | 6 | 3.19% |
|  |  | 40.96% |  |  |  |
| Female | 77 |  | 12 - 17 years | 24 | 12.77% |
| BMI < 18 | 13 | 6.91% | 18 - 39 years | 100 | 53.19% |
| 18 ≤ BMI < 24 | 113 | 60.11% | 40 - 59 years | 50 | 26.60% |
| BMI ≥ 24 | 62 | 32.98% | > 60 years | 8 | 4.26% |

Table 2 shows the distribution of services, and PER is primarily concentrated in the Neurology Outpatient Department, with a total of 724 prescriptions, accounting for 87.12% of cases.

**Table 2.** Distribution of Department and Diagnosis for the use of PER

| Department | Copies | Proportion |
| --- | --- | --- |
| Neurology clinic | 724 | 87.12% |
| Epilepsy clinic | 102 | 12.27% |
| Thoracic surgery outpatient I | 2 | 0.24% |
| Pulmonary nodule specialist | 1 | 0.12% |
| Neurosurgery clinic | 1 | 0.12% |
| Pediatric Neurology Clinic | 1 | 0.12% |
| Total | 831 | 100% |

Table 3 shows the diagnosis for the prescriptions, and it can be seen that the PER is primarily used for epilepsy, symptomatic epilepsy [secondary epilepsy], epileptic seizures, and other related disorders. More information about diagnosing PER can also be found in Table 3.

**Table 3.** Diagnosis of prescription diseases of PER tablets

| Disease diagnosis | Copies | Proportion |
| --- | --- | --- |
| Epilepsy | 555 | 66.79% |
| Symptomatic epilepsy [Secondary epilepsy] | 99 | 11.91% |
| Symptomatic epilepsy | 19 | 2.29% |
| Pain | 15 | 1.81% |
| Intracranial space-occupying lesion (post glioma surgery) | 11 | 1.32% |
| Post-traumatic syndrome, epilepsy | 13 | 1.56% |
| Juvenile absence epilepsy | 10 | 1.20% |
| Dizziness, headache syndrome, other specified | 13 | 1.56% |
| Burn | 9 | 1.08% |
| AVM embolization postoperative | 8 | 0.96% |
| Encephalitis (Autoimmune encephalitis) | 8 | 0.96% |
| Epilepsy (Psychomotor seizure) | 6 | 0.72% |
| Seizures, intracranial parasitic infection (after surgery) | 6 | 0.72% |
| Brain abscess (Postoperative) | 6 | 0.72% |
| Sjögren 's Syndrome\[Sjogren' s] | 6 | 0.72% |
| Limbic encephalitis (?) | 5 | 0.60% |
| Observation of Suspected Diseases and Conditions | 6 | 0.72% |
| Diffuse astroglioma WHO Grade II postoperative | 3 | 0.36% |
| Lumbar disc herniation | 3 | 0.36% |
| Epilepsy, mesial temporal lobe epilepsy with hippocampal sclerosis | 2 | 0.24% |
| Antisynthetase syndrome | 1 | 0.12% |
| Meningioma | 1 | 0.12% |
| Partial seizures | 1 | 0.12% |
| Frontal lobe epilepsy | 1 | 0.12% |
| Anxiety state | 1 | 0.12% |
| Immune-mediated encephalitis | 1 | 0.12% |
| Hydrocephalus | 1 | 0.12% |
| Postoperative chemotherapy for malignant tumor | 1 | 0.12% |
| Sleep disorder | 1 | 0.12% |
| Other (hepatitis B, sleep disorder, diabetes mellitus, bradycardia, chronic gastritis, methemoglobinemia) | 19 | 2.29% |
| Total | 831 | 100.00% |

Table 4 details the usage of PER within FAHXU (compared to the drug instructions). Overall, 119 prescriptions, 14.32%, were inconsistent with the indications for the PER; 56 prescriptions with inappropriate dosing frequency, representing 6.74%; three prescriptions with inconsistent dosage, 0.36% repentance, and six prescriptions had inappropriate dosage interval adjustments, 0.72%. More information on the use of the PER within FAHXU could be found in Table 4 as well.

**Table 4.**
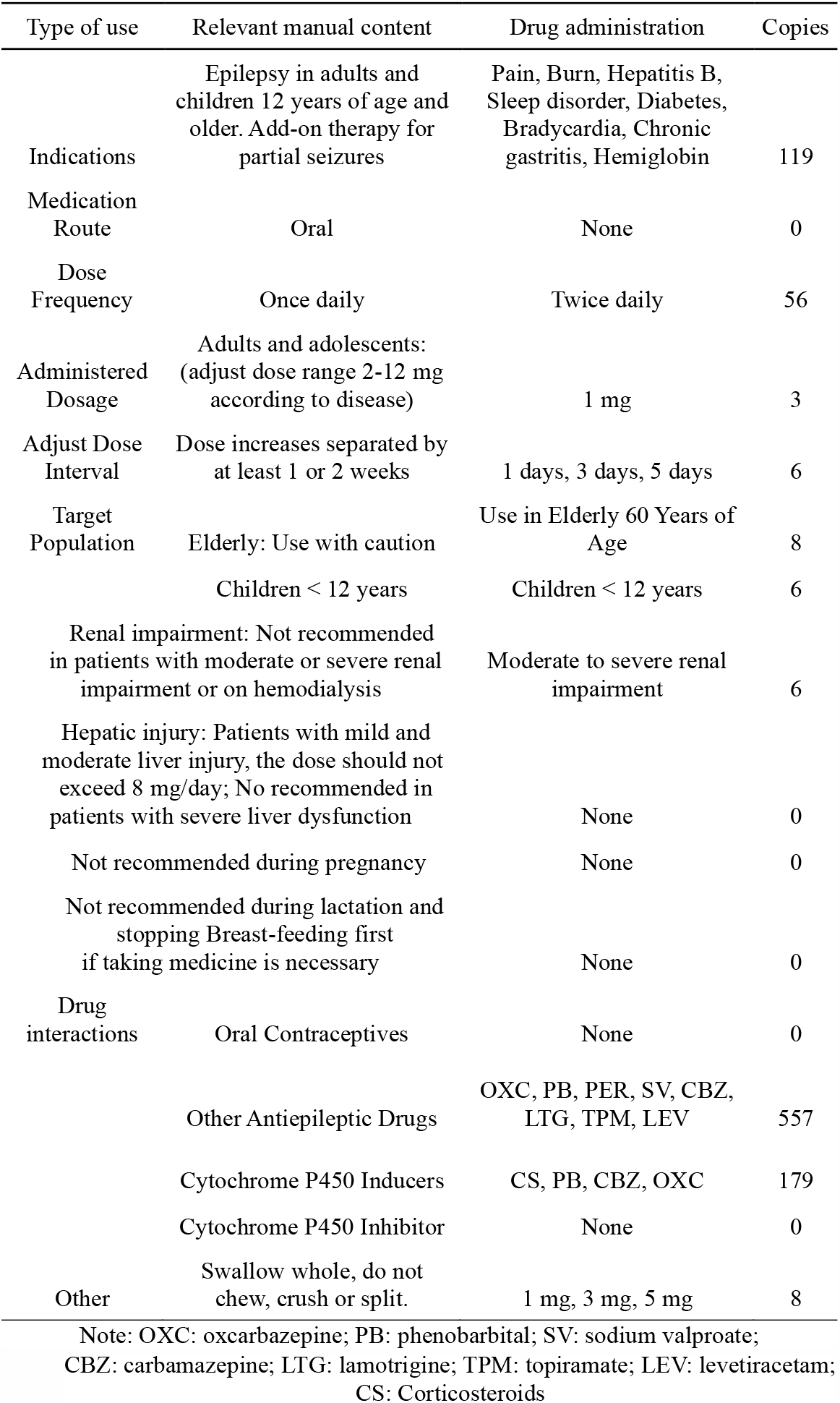
Administration of PER Tablets (as compared to the package insert)

Table 5 shows the combination use of PER in FAHXU hospital, with 311 prescriptions were combination with one antiepileptic drug, 231 prescriptions were in combination with two antiepileptic drugs, and 15 prescriptions were combination with three antiepileptic drugs.

**Table 5.** Concomitant medication of PER Tablets

| Concomitant medication | Drug Name | Combined Antiepileptic Drug Varieties | Copies |
| --- | --- | --- | --- |
| Novel antiepileptic drug | LEV (378), SV (296), OXC (81), TPM (67), LTG (61) | Concomitant use of an antiepileptic drug | 311 |
| Traditional antiepileptic drugs | CBZ (32), PHB (25) | Combined with two antiepileptic drugs | 231 |
| Sedative hypnotic drugs | CZP (66), AL (6), ZBT (3), DP (1) | Combined with three antiepileptic drugs | 15 |
| Vitamins & Electrolytes | Vitamin B1 (125), POT CL (1), FA (1) |  |  |
| Antidepressants | Duloxetine (12), Lexapro (9) |  |  |
| Hormones | PTA (10) |  |  |
| Gastrointestinal system medication | OMP (9) |  |  |
| Hepatoprotective drugs | PPC (6) |  |  |
| Antipsychotics | Olanzapine (6) |  |  |
| Antimigraine drugs | Flunarizine (2) |  |  |
| Central stimulant | Piracetam (4) |  |  |
| Others | Botox A (5) |  |  |

Note: AL: alprazolam; CBZ: Carbamazepine; PHB: Phenobarbital; DP: diazepam; ZBT: zolpidem; CZP: Clonazepam; LEV: Levetiracetam; LTG: lamotrigine; FA: folic acid; POT CL: potassium chloride; Lexapro (Escitalopram); PTA: Prednisone acetate; OMP: Omeprazole; OXC: oxcarbazepine; PPC: Polyene phosphatidylcholine; SV: sodium valproate; TPM: topiramate; Botox A: Botulinum toxin A;

## Discussion

### Analysis of the Basic Characteristics of Patients Using PER

As shown in the table 1, 111 male patients, accounting for 59.04%, and 77 female patients, accounting for 40.96%, were treated with PER. This is consistent with fact that there are more males and fewer females with epilepsy in China. Patients under the age of 12 accounted for 3.19%, and according to the medication instructions, “the safety and efficacy of PER in children under 12 years old has not yet been established.” The efficacy and safety of PER in 96 Chinese pediatric patients aged 2-14 years were reported by Qu and Chen, et al^18^. Treatment effect was good, safety was high and adherence was good. PER has been on the commercially available since 2008 and has been approved by the U.S. FDA in 2012 for the treatment of focal seizures in patients 12 years and older^19, 20^, with progressively expanding indications in a variety of countries. PER is a new indication that was approved in China in July 2021, and can be used to treat patients with partial-onset seizures (with or without secondary generalized seizures) in adults and children aged 4 years and older^21^. Pending the author’s investigation and analysis, the instructions have not been updated in time. Table 1 shows that, 32.98% of patients had a BMI ≥ 24. Studies have shown that the use of antiepileptic drugs can cause weight gain. It is possible that the obese population in this study is related to PER use. Due to the short duration of clinical use of PER in China, there is a need to study the side effects leading to obesity.

### Analysis of medication of PER in FAHXU hospital

#### Indications for PER

According to Table 3, PER is primarily used for the treatment of epilepsy, symptomatic epilepsy [secondary epilepsy], epileptiform convulsions and other diseases. Its indication is in accordance with the package insert. The indications include alternative diagnoses such as traumatic brain injuries and encephalitis, which are not included in the medication labeling, but these conditions may cause seizures. Traumatic brain injury (TBI) is a common cause of epilepsy, epidemiologic studies have demonstrated a clear relationship between the severity of injury and the likelihood of developing epilepsy ^22, 23^. In addition, encephalitis, an inflammation and swelling of the brain, is a leading cause of acute symptomatic seizures and subsequent epilepsy, with approximately one-third of patients presenting with seizures^24, 25^. Encephalitis may be of either infective or autoimmune in origin^26^. We also included patients with intracranial space-occupying lesions, seizure symptoms have been reported in 30-50% of patients with brain tumors, especially benign slow-growing brain tumors^27-29^. Even if the pain diagnosis does not match the routine use of antiepileptics, after the onset of disease, patients with epilepsy are subject to pain or headache^30-32^. Studies have shown that during epileptic seizures, nerves in the brain are over discharged, leading to local tissue disruption, abnormal blood supply, and ultimately physical deterioration and death of some cells, resulting in the development of pain symptoms^33^. The above analysis suggests that there is inappropriate prescriptions use of PER in FAHXU hospital. Consulting the cases reveals that most of them were seizures caused by other conditions, because physicians did not fully diagnose epilepsy, the indications were not suitable.

### Frequency of dosing with PER

Table 4 shows the most common inappropriate dosing frequency is twice daily (56 cases, 6.74%). The pharmacokinetic studies of PER have shown that peak concentration is reached approximately one hour after oral administration, with an average half-life of 105 hours. The elimination half-lives after single and multiple doses are 52-129 hours and 66-90 hours, respectively. Steady-state blood concentrations are reached after 14 days of multiple doses. Pharmacokinetic characteristics are consistent with a single-compartment model of first-order elimination. As a result, a once-daily dose is appropriate. PER is primarily used to treat epilepsy by suppressing abnormal nerve discharge in the brain. Seizures are most likely to occur while sleeping, when the inhibitory effect of the cerebral cortex is reduced, facilitating epileptic foci that have lost inhibition to produce abnormal discharges. Furthermore, neurons are also more likely to exhibit synchronized discharges, which can trigger epileptic seizures, during light sleep. Taking drugs at night can effectively control abnormal synchronized discharges of neurons, in addition to improving seizure status, alleviating the condition, and promoting early recovery from the disease. Therefore, it is recommended to take PER at night.

### Concomitant use of PER

Pharmacotherapy is the mainstay of treatment for epilepsy. Relevant guidelines suggest beginning with low-dose monotherapy. If monotherapy fails to control seizures or side effects occur, another type of monotherapy may be considered. PER has a different mechanism than other antiepileptic drugs. It is often treated as an adjunctive therapy to other antiepileptics. This survey shows that it is usually used in combination with other medications, with a total of 331 prescriptions for combination with one antiepileptic drug, 231 prescriptions for combination with two antiepileptic drugs, and 15 prescriptions for combination with three antiepileptic drugs. However, when used in combination with other antiepileptic drugs, attention should be given to drug-drug interactions. Antiepileptic drugs such as carbamazepine, phenytoin, and oxcarbazepine, which concomitantly take enzymatic inducers in the liver, have been shown to be effective, may enhance the rate of clearance of this product, leading to a reduction in the plasma concentration of PER^14^. Consequently, when combined with such antiepileptic drugs, attention must be paid to titrating the drug dosage to achieve efficacy.

Relatively high proportions of vitamin B1 is also prescribed, in addition to antiepileptic drugs. Vitamin B1 belong to the group of nutritional neurotrophic vitamins and have a certain effect of improvement in patients with epilepsy. They can stabilize the nerves and have a sedative effect. The combination of PER and VB1 can have a positive effect on the severity and frequency of seizures and on the side effects of the drugs on the body. It has been demonstrated by studies: Patients with epilepsy are often associated with the development of mental illness. ^34-36^. Depression is the most common mental illness in these patients, with an incidence as high as 30%, which is 5 ∼ 20 times that of the general population. Patients with epilepsy have a prognosis that is influenced by the comorbidity of mental illness, resulting in poor quality of life and early death^36-39^. Botulinum toxin type A, Botulinum Neurotoxin (BoNT) is also a drug combination. It inhibits acetylcholine release at axon terminals in surrounding neurons, paralyzes flaccid muscle, and reduces glandular secretions. BoNT has been empirically used in a wide variety of ophthalmological, gastrointestinal, urologic, orthopedic, dermatologic, secretory and pain disorders^40, 41^. Research results show BoNT can inhibit neuronal excitability, reducing acute seizures as well as chronic spontaneous epileptic seizures. Thereby reducing neuronal damage and abnormal brain firing, providing a novel means of treating epilepsy^42-44^.

### Characteristics of PER

During this investigation, it was discovered that PER tablet had been ruptured and the drug dosage was 1 mg, 3 mg and 5 mg. The PER used in FAHXU is a scarless tablet made by Eisai Europe limited and is an unmarked tablet. It is clear from the instructions that it should be swallowed whole and should not be chewed, crushed, or divided. PER tablet scission can lead to inaccurate dosing and debris creation. Although the loss of some debris may have little effect on the therapeutic effect in short term, antiepileptic drugs often require long-term use. Thus, drugs should be used in strict accordance with medication instructions.

### Dose Adjustment Interval for PER

The study showed that some patients adjust the medication dosage within a range of 1 day to 16 weeks. The instruction of PER indicated that the dose should be adjusted at intervals of 2 weeks or at least one week. Studies have shown that the development of drug tolerance can be delayed by a reasonable interval between doses^45^. Tolerance to antiepileptic drugs is an important cause of clinical treatment failure^46^. Therefore, medication dosage adjustments should be made according to the drug instructions in a regular and reasonable manner, in order to slow down the development of drug tolerance.

## Conclusions

In conclusion, epilepsy is a chronic neurologic disorder that severely affects patients’ daily lives. Not only a medical problem, but also an important public health and social issue. Pharmacological is the most common method of treating epilepsy, and the clinical demand for antiepileptic drugs is increasing as a result of the long course of epilepsy and the prolonged duration treatment. The novel antiepileptic drug Perampanel offers a new option for the treatment of epilepsy. Irrational use of PER still occurs in FAHXU hospital. Changes in patients’ illnesses and adverse event after medication should receive more attention. It is a need for an appropriate medication use assessment system to strengthen supervision of PER clinical use and promote rational medication use.

## Data Availability

All data produced in the present study are available upon reasonable request to the authors.

## Abbreviations

AED: antiepileptic drug
AL: alprazolam
Botox A: Botulinum toxin A
CBZ: Carbamazepine
CS: Corticosteroids
CZP: Clonazepam
DP: diazepam
FA: folic acid
LEV: Levetiracetam
LEXAPRO: Lexapro (Escitalopram)
LTG: lamotrigine
OMP: Omeprazole
OXC: oxcarbazepine
PB: phenobarbital
PHB: Phenobarbital
PTA: Prednisone acetate
PPC: Polyene phosphatidylcholine
POT CL: potassium chloride
SV: sodium valproate
TPM: topiramate
ZBT: zolpidem

## Acknowledgements

The authors would like to acknowledge the assistance of staff at the Department of Information Technology at the hospital for data extraction.

## Author Contributions

LL and YH contributed to data collection, data analysis and writing of the article. ZS, LM gave advice on the design of the study. LL and YH helped collect data. All authors revised the manuscript and eventually approved it for publication. LL was responsible for the overall content as guarantor who accepted full responsibility for the finished work and the conduct of the study, had access to the data, and controlled the decision to publish.

## Funding

The authors have not declared a specific grant for this research from any funding agency in the public, commercial or not-for-profit sectors.

## Competing interests

None declared.

## Patient and public involvement

Patients and/or the public were not involved in the design, or conduct, or reporting, or dissemination plans of this research.

## Patient consent for publication

Not applicable.

## Ethics approval

This study involves human participants and was approved by the Ethics Committee of the First Affiliated Hospital of Xiamen University.

### Provenance and peer review

Not commissioned; externally peer reviewed.

### Data availability statement

Data are available upon reasonable request.

## Open access

This is an open access article distributed in accordance with the Creative Commons Attribution Non-Commercial (CC BY-NC 4.0) license, which permits others to distribute, remix, adapt, build upon this work non-commercially, and license their derivative works on different terms, provided the original work is properly cited, appropriate credit is given, any changes made indicated, and the use is non-commercial. See: http://creativecommons.org/licenses/by-nc/4.0/.

## Notes

### Competing Interest Statement

The authors have declared no competing interest.

### Funding Statement

This study did not receive any funding.

